# Deciphering the Effect of High-Density Lipoprotein Cholesterol on Renal Function: a Mendelian Randomization Study

**DOI:** 10.1101/2023.10.18.23297196

**Authors:** Nhu Ngoc Le, Tran Quoc Bao Tran, Dipender Gill, Sandosh Padmanabhan

## Abstract

**Background:** The causal relationship between high-density lipoprotein cholesterol (HDL) and cardiovascular protection remains unresolved. Mounting evidence now points towards a link between lipid levels, including HDL, and renal function. However, previous epidemiological and animal studies provide a mixed picture. This study investigates the causal interplay between HDL and renal function by leveraging the specific and substantial increase in HDL achieved with cholesteryl ester transfer protein (CETP) inhibition.

**Method and Results:** Two-sample and multivariable Mendelian randomization (MR) methods were used to explore the causal impact of HDL through genetically-proxied CETP inhibition on serum cystatin C-estimated glomerular filtration rate (eGFRcys). Secondary analyses delved into other renal markers and systolic blood pressure (SBP). Summary-based Mendelian Randomization (SMR) and genetic colocalisation analyses were used to evaluate the probability of shared causal variants within a 100Kb window of the gene.

Genetically-proxied CETP inhibition, using HDL level as a biomarker, was linked to lower eGFRcys (effect size per 1 SD increase in HDL, −0.008, 95% CI −0.011 to - 0.005; p = 1.38 × 10^−06^) and reduced chronic kidney disease (CKD) risk (OR 0.895 [0.838, 0.956]; p = 0.001). The HDL-eGFRcys relationship persisted after adjusting for low-density lipoprotein cholesterol (LDL) and SBP in multivariable MR, but the association with CKD risk attenuated. Decreased CETP expression in blood was associated with lower eGFRcys (effect size per 1-SD, −0.008 [−0.016, −0.001]; p_SMR_ = 0.029), a reduced CKD risk (OR, 0.85 [0.74, 0.98]; p_SMR_ = 0.03), and lower SBP (−0.71 [−1.177, −0.244]; p_SMR_ = 0.003). Colocalisation results indicated low posterior probabilities for both shared and distinct causal variants between CETP gene expression and eGFRcys.

**Conclusion:** MR analyses support a causal inverse relationship between HDL and eGFRcys that is independent of SBP. The results warrant further studies to validate the nuanced roles of HDL and LDL on renal function.

## 1. Introduction

High-density lipoprotein cholesterol (HDL) is conventionally understood to protect against cardiovascular diseases (CVD) through mechanisms like reverse cholesterol transport and anti-inflammatory activities. Observational studies have consistently reported an inverse association between HDL levels and the risk of coronary heart disease (CHD). Yet, its causal role in the etiology of CHD remains unresolved (1–3).

Pharmacological inhibition of cholesteryl ester transfer protein (CETP) has emerged as a promising therapeutic strategy for modulating HDL. Such inhibition has been shown to substantially increase HDL levels while decreasing non-HDL cholesterol, particularly low-density lipoprotein cholesterol (LDL). Despite setbacks in early clinical trials, the REVEAL trial demonstrated potential cardiovascular benefits from CETP inhibition, largely attributed to the reduction in non-HDL cholesterol levels (4). Concurrently, some CETP inhibitors such as torcetrapib and anacetrapib have been associated with modest increases in systolic blood pressure (SBP) in all Phase III randomized controlled trials (RCTs), and this increase appears to correlate with the degree of elevation in HDL levels and has been attributed to off-target effects of the drug (4–6).

Moreover, there is a burgeoning body of evidence implicating lipid levels, including low HDL, in the decline of renal function and the onset and progression of chronic kidney disease (CKD). Animal models strongly suggest that dyslipidemia could be both a driver and a marker for renal damage (7). Epidemiological studies, however, present a convoluted picture, with inconsistent associations between lipid levels and renal function across different populations (8–14). Causal inference through Mendelian randomisation studies for HDL cholesterol and renal function have supported either no effect of HDL on eGFR in European (15) and African ancestries (16), a decrease in eGFR but a decrease in risk of CKD with an increase in HDL (17, 18) or an increase in eGFR with an increase in HDL (19), in contrast to observational studies.

A substantial limitation in existing research is the use of serum creatinine-based estimated glomerular filtration rate (eGFR) to gauge renal function. This measure is influenced by various factors like muscle mass, diet, and age (20, 21). Cystatin C offers a more reliable alternative, being less susceptible to individual patient characteristics and thought to be a more sensitive marker of renal function (22, 23).

The confounding effect of SBP increase consistently associated with CETP inhibition in clinical trials and the established impact of high SBP on renal dysfunction underscore the need for a more nuanced investigation into the role of HDL on renal function. Given these complexities and gaps in understanding, our study aims to elucidate the causal relationship between HDL and renal function by leveraging the specific and substantial increase in HDL achieved with CETP inhibition which can be proxied using genetic variants in the CETP gene. We employ a Mendelian randomization approach to investigate the causal association between genetically proxied CETP inhibition and various traits of renal function including cystatin-based eGFR (eGFRcys).

## 2. Method

All data used in this study are publicly available. Relevant ethical approval and patient consent were obtained in the contributing studies.

### Study Design

The study design is illustrated in **Figure 1**. Two-sample Mendelian randomization (MR) analysis was conducted to investigate the causal effect of genetically proxied CETP inhibition on glomerular filtration rate estimated by serum cystatin C (eGFRcys) as a primary outcome. We also conducted the analysis on glomerular filtration rate estimated by creatinine (eGFRcrea), blood urea nitrogen (BUN), urine albumin-to-creatine (uACR), microalbuminuria, chronic kidney disease (CKD) and systolic blood pressure (SBP). The potential mediating pathways of CETP inhibition on the outcomes were further explored through a joint modelling of LDL, HDL, and SBP using multivariable MR analysis. The causal association between genetically proxied lipids (HDL, LDL levels) more generally and the outcomes was investigated using genetic variants associated with the lipid traits from the whole genome with and without the variants located within 1 Mb of the CETP gene. Moreover, drug target MR analysis of 3-hydroxy-3-methylglutaryl-coenzyme A reductase (HMGCR) on the same outcomes was performed to differentiate the anticipated effects predominantly HDL effect of CETP versus the predominantly LDL effects of HMGCR inhibition.

**Figure 1.**
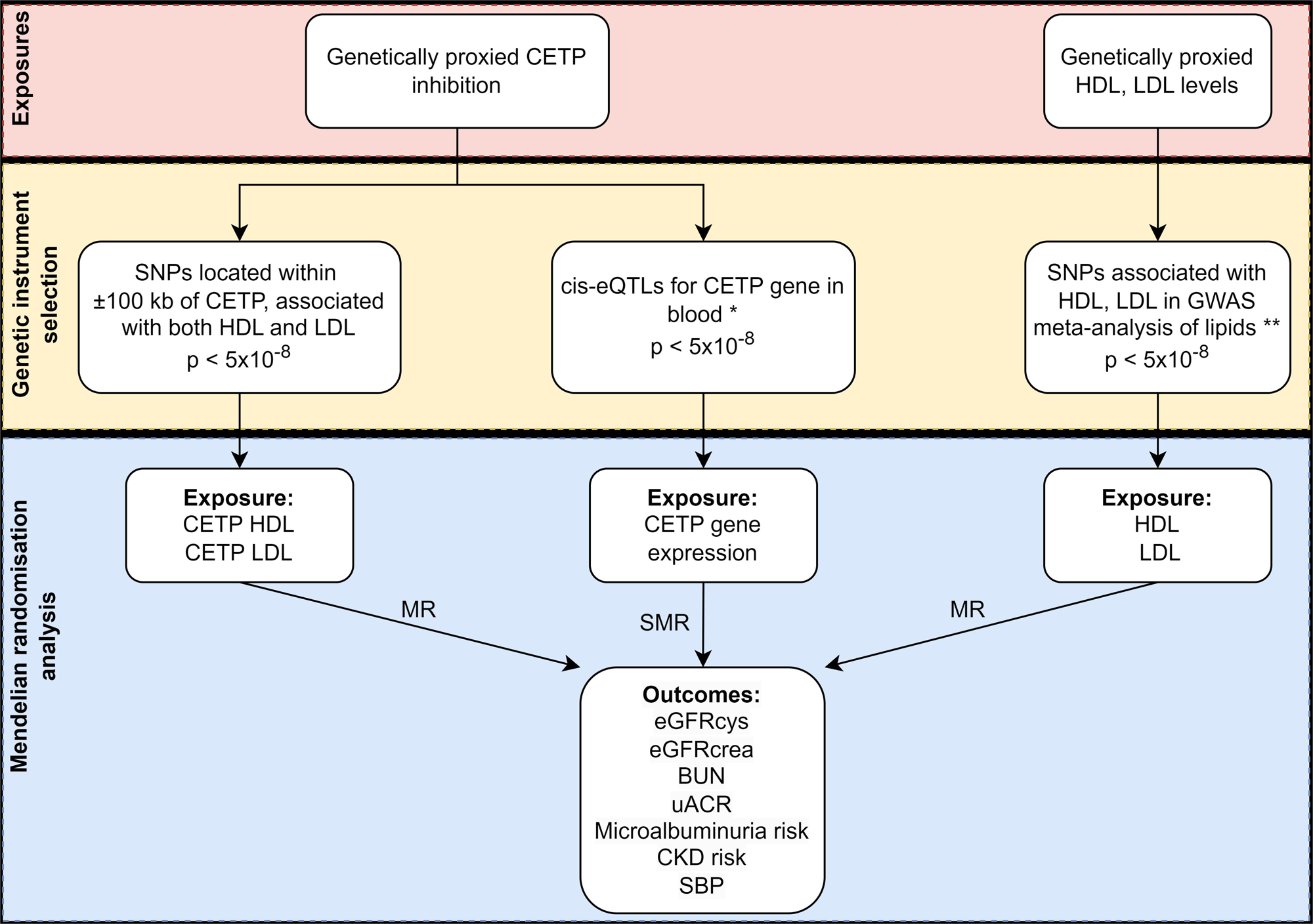
Flowchart depicting the study design. CETP indicates cholesteryl ester transfer protein; LDL, low-density lipoprotein; HDL, high-density lipoprotein; SNP, single nucleotide polymorphism; eQTL, expression quantitative trait loci; GWAS, genome-wide association study; MR, Mendelian randomization; SMR, summary-based Mendelian randomization; eGFRcys, serum cystatin C-estimated glomerular filtration rate; eGFRcrea, glomerular filtration rate estimated by creatinine; BUN, blood urea nitrogen; uACR, urine albumin-to-creatinine ratio; MA, microalbuminuria; CKD, chronic kidney disease; (*), eQTLs were extracted from the eQTLGen consortium (30); (**), lipids SNPs were extracted from the Global Lipids Genetics Consortium GWAS meta-analysis (28).

For drug MR analysis, we applied two different approaches to select genetic variants to proxy perturbation of the drug target: (i) select variants that are located within or near the CETP gene (cis-variants) and are strongly associated with biomarkers (HDL and LDL levels) known to be affected by CETP inhibition; (ii) select cis-variants that are associated with the CETP gene expression in blood (expression quantitative trait loci or eQTLs). We used genetic associations with HDL level as the primary biomarker to scale MR estimates, with secondary analyses considering genetic associations with LDL instead of HDL.

### Two-sample MR

The MR approach uses genetic variants as instrumental variables to investigate the causal effect of an exposure on an outcome. By leveraging the random allocation of genetic variants at conception, this method can mitigate biases arising from confounding factors and reverse causation, which are fundamental limitations in observational studies (24). MR can be extended to investigate a causal effect of drug target perturbation on outcomes by using genetic variants related to the function or expression of the gene encoding the drug target (25, 26). Summary-based Mendelian Randomization (SMR), an extension of the MR method, integrates summary-level data from independent GWAS and eQTL studies to investigate the causal association between gene expression and an outcome of interest (27).

### Genetic Instrument Selection

#### Genetic proxies for lipid-lowering drug target

To proxy the effect of lipid-lowering drug targets (CETP, HMGCR), genetic instruments were identified as uncorrelated (r^2^ < 0.1) single nucleotide polymorphisms (SNPs) (i) located within 100 kb on either side of the gene (CETP, chromosome 16: 56,995,762-57,017,757; HMGCR, chromosome 5: 74,632,154-74,657,929, on build GRCh37/hg19), (ii) and associated with biomarkers (HDL and LDL levels for CETP, LDL for HMGCR) at the genome-wide significance level (p-value of 5 × 10^−08^) in the Global Lipids Genetics Consortium (GLGC) GWAS meta-analysis of 1,320,016 European-ancestry individuals (28). The summary statistics data are on the scale of a 1-SD change in the levels of LDL/HDL in blood for each additional effect allele. F-statistics were calculated to estimate the instrument strength (29).

#### Genetic proxies for gene expression

Genetic proxies for the drug target expression were identified as SNPs that were associated with the expression of the genes in the blood (minor allele frequency > 1%, eQTL p-value < 5 × 10^−08^), using summary statistics data from the eQTLGen consortium (n = 31,684) (30). Only cis-eQTLs (eQTLs located within a 100kb window of the gene) were included as the instrumental variables for the analysis. The beta coefficient represents the SD change in the expression level of the gene per copy of the effect allele (30). F-statistic was calculated to assess the strength of the instrument (29).

For the validation of genetic instruments, SMR analysis was performed to test whether changes in gene expression in blood reflect a lipids-lowering association with the drug targets (increased HDL and decreased LDL levels for CETP inhibition and decreased LDL levels for HMGCR inhibition). The effect size estimates from SMR indicate the SD changes in LDL or HDL per 1-SD increase in gene expression.

#### Genetic proxies for LDL, HDL levels

To investigate the causal association between the lipid traits (LDL, HDL levels) and the outcomes, genetic proxies for the lipid trait (LDL or HDL levels) were identified as SNPs that associated with the lipid trait at the genome-wide significance level (p-value < 5 × 10^−08^) across the entire genome. Due to a larger number of SNPs identified from the whole genome, a clumping threshold of r^2^ < 0.001 was used to reduce the bias arising from linkage disequilibrium.

### Outcome Data

Genetic association estimates for eGFRcys, eGFRcrea, and BUN were obtained from the Chronic Kidney Disease Genetics (CKDGen) Consortium and UK Biobank (UKB) GWAS meta-analysis of 460,826 (eGFRcys), 1,004,040 (eGFRcrea), and 852,678 (BUN) individuals, predominantly European ancestry (31). eGFRcys was estimated from cystatin C measurements using either the formula by Steven et al. (32) (CKDGen) or the Chronic Kidney Disease Epidemiology Collaboration (CKD-EPI) formula (33) (UKB). eGFRcrea was obtained from serum creatinine based on the CKD-EPI (UKB, CKDGen for individuals over 18 years of age) or the Schwartz (34) (CKDGen for individuals 18 years of age or younger) formula. Both eGFRcys, and eGFRcrea were log-transformed using a natural logarithm. Blood urea measurements, expressed in mg/dL, were multiplied by 2.8 to derive BUN values, which were subsequently log-transformed using a natural logarithm (31). Genetic association estimates for uACR and microalbuminuria risk were extracted from a GWAS meta-analysis of 547,361 individuals (European ancestry) and 348,954 individuals (> 96% European ancestry), respectively (35). UACR was assessed in mg/g, calculated as urinary albumin (mg/l) to urinary creatinine (mg/dl), multiplied by 100. Microalbuminuria cases were defined as uACR greater than 30, while controls were identified as uACR less than 10 mg/g (35). Genetic association estimates for CKD were obtained from the Chronic Kidney Disease Genetics (CKDGen) Consortium GWAS meta-analysis (41,395 cases and 439,303 controls of European ancestry) (36). CKD was characterised as an eGFRcrea < 60 ml min^−1^ per 1.73 m^2^ (36). Genetic association estimates for SBP were derived from a GWAS meta-analysis of 757,601 European-ancestry individuals from the UKB and the International Consortium of Blood Pressure (ICPP) (37). The characteristics of the population and phenotype definitions were described elsewhere (31, 35–37).

### Statistical Analysis

MR requires three main instrumental variable assumptions: firstly, the genetic variant is strongly associated with the exposure; secondly, the genetic variant has no common cause with the outcome; and thirdly, the genetic variant only affects the outcome via the exposure (38).

For each of the genetic instruments used in MR analysis, the genetic associations with the exposure and outcome were harmonized by aligning effect alleles. No exclusions were made for palindromic variants. To maintain consistency in the genetic instruments used across various analyses, proxies were not used. Inverse-variant weighted (IVW) MR was conducted as the primary analysis to estimate the causal effect of genetically proxied CETP inhibition on the outcomes. MR estimates were presented as beta with a 95% confidence interval (95% CI) orientated to the drug target effect direction: lower LDL level, higher HDL level, and lower gene expression level.

The simple median, weighted median and MR Egger were used in the sensitivity analysis to examine the robustness of the results to potential pleiotropic effects of the variants (39, 40). The median-based methods are more robust to genetic variants with strongly outlying causal estimates than the IVW method. These methods calculate the median of the ratio instrumental variable estimates evaluated using each genetic variant. The simple median method calculates the median value without considering the weight of each variant. It can provide a consistent MR estimate of the causal effect when at least 50% of the genetic variants are valid.

Whereas the weighted median method incorporates the weight of each genetic variant when calculating the median value and can give a consistent MR causal estimate if at least 50% of the weight comes from valid instrumental variables (39). The MR Egger method can test for the presence of horizontal pleiotropy by its intercept test (40). Horizontal pleiotropy occurs when genetic variants affect outcome not only through the exposure, leading to the violation of the exclusion restriction assumption in MR analysis (41). The MR Egger method gives a valid MR estimate if any direct effect of the genetic instruments on the outcome is not correlated with their associations with the exposure (40).

Multivariable Mendelian randomisation analysis (MVMR) (42) was conducted to explore potential mediating pathways of CETP inhibition effect on the outcomes through a joint modelling of HDL, LDL, and SBP. Cis-variants (SNPs located within a 100kb window of the gene) associated with either HDL, LDL, or SBP at genome-wide significance were combined and clumped to pair-wise LD r^2^ < 0.1 after ordering by lowest p-value. For each of the instruments, the genetic association estimates with the exposure, the mediator, and the outcome were harmonized by aligning effect alleles.

IVW, simple median, weighted median, MR Egger, and MVMR analyses were performed using the MendelianRandomization package (version 0.6.0) in R. LD clumping was conducted using the TwoSampleMR package in R (43); LD among the SNPs was estimated based on the 1000 Genomes European reference panel (44).

#### Summary-based Mendelian randomization (SMR)

SMR was performed to test if the drug target gene expression levels were associated with outcomes of interest (27). Cis-variants associated with blood gene expression (cis-eQTLs) were used to proxy the effects of CETP inhibition (exposure) on LDL and HDL levels (positive controls), on eGFRcys (primary outcome), eGFRcrea, BUN, uACR, microalbuminuria, CKD, and SBP (secondary outcomes). SMR estimates were presented as beta per 1-SD change in blood CETP gene expression, with the direction of gene expression change being orientated to reflect an LDL-lowering / HDL-increasing association. The heterogeneity in dependent instruments (HEIDI) test is performed to explore if the association between gene expression and the outcome is due to a linkage scenario (SNP that affects gene expression is in high LD with another SNP that affects the outcome) instead of true causality (27). A HEIDI p-value < 0.01 indicates that the observed association is most likely due to the linkage scenario. eQTLs in very high or low LD (r^2^ > 0.9 or r^2^ < 0.05) with the top associated eQTL were excluded in the HEIDI test to avoid the issue of collinearity (27). SMR version 1.03 was used to conduct all SMR analyses and HEIDI tests (27).

#### Colocalisation analysis

For any significant association between the gene expression and the outcome, genetic colocalisation analyses were conducted to estimate the posterior probability for a common causal variant between the two traits, within a 100Kb window of the genes. Coloc v5.2.1 R package (45) was used to perform colocalisation analysis. Coloc conducted a Bayesian test for colocalisation in a genomic locus between two traits, assuming at most one causal variant per trait in the genomic region. Coloc calculates five posterior probabilities for the competing models:

H_0_: No variant associated with either trait in the region
H_1_: A variant associated with trait 1, but not trait 2 in the region
H_2_: A variant associated with trait 2, but not trait 1 in the region
H_3_: Distinct causal variants for trait 1 and trait 2 in the region
H_4_: A shared causal variant for both traits in the region

Default priors were used for the analysis: p_1_ = 10^−4^, p_2_ = 10^−4^, p_12_ = 10^−5^ (p_1_ is the prior probability for a variant being associated with trait 1, p_2_ is the prior probability for a variant being associated with trait 2, p_12_ is the prior probability a variant being associated with both traits).

To account for the risk of Type I error due to multiple comparisons, a stringent multiple testing correction was applied. Given that 14 primary analyses were performed (7 outcomes and 2 primary exposures - genetically proxied CETP inhibition using HDL as biomarker and genetically proxied HDL), a Bonferroni correction was utilized to adjust the significance level. Consequently, a p-value of less than 0.0036 (0.05/14) was considered statistically significant to ensure the robustness of the findings against the possibility of false positives arising from multiple testing.

## 3. Results

### Mendelian Randomization Analyses

#### Mendelian randomization of HDL and LDL changes through genetically proxied CETP inhibition

Within the CETP locus, ten uncorrelated SNPs (r^2^ < 0.1) were identified as genetic proxies for CETP inhibition (**Table S1**). All selected SNPs have F-statistics greater than 10, indicating a low possibility for bias due to weak instruments. Considering the genetic association with HDL level as the exposure, genetically proxied CETP inhibition was associated with lower eGFRcys (effect size per 1 SD increase in HDL level, −0.008, 95% CI −0.011, −0.005; p = 1.38 × 10^−06^) (**Figure 2, Figure S1, Table S2).** The MR estimates obtained from the simple median, weighted median and MR-Egger were consistent with the estimate from the IVW method. The MR-Egger intercept test found no evidence of directional pleiotropy (intercept p-value 0.25) (**Table S2**). Additionally, genetically proxied CETP inhibition (reflected as an increase in HDL level as exposure) was significantly associated with a decreased risk of CKD (OR per 1-SD increase in HDL level, 0.895, 95% CI, 0.838, 0.956; p = 0.001), nominally associated with lower SBP (effect size, −0.531, 95% CI −0.915, - 0.146) (**Figure 3**), but not associated with eGFRcrea, BUN, uACR, and MA risk (**Table S2, Figure S2-6**).

**Figure 2.**
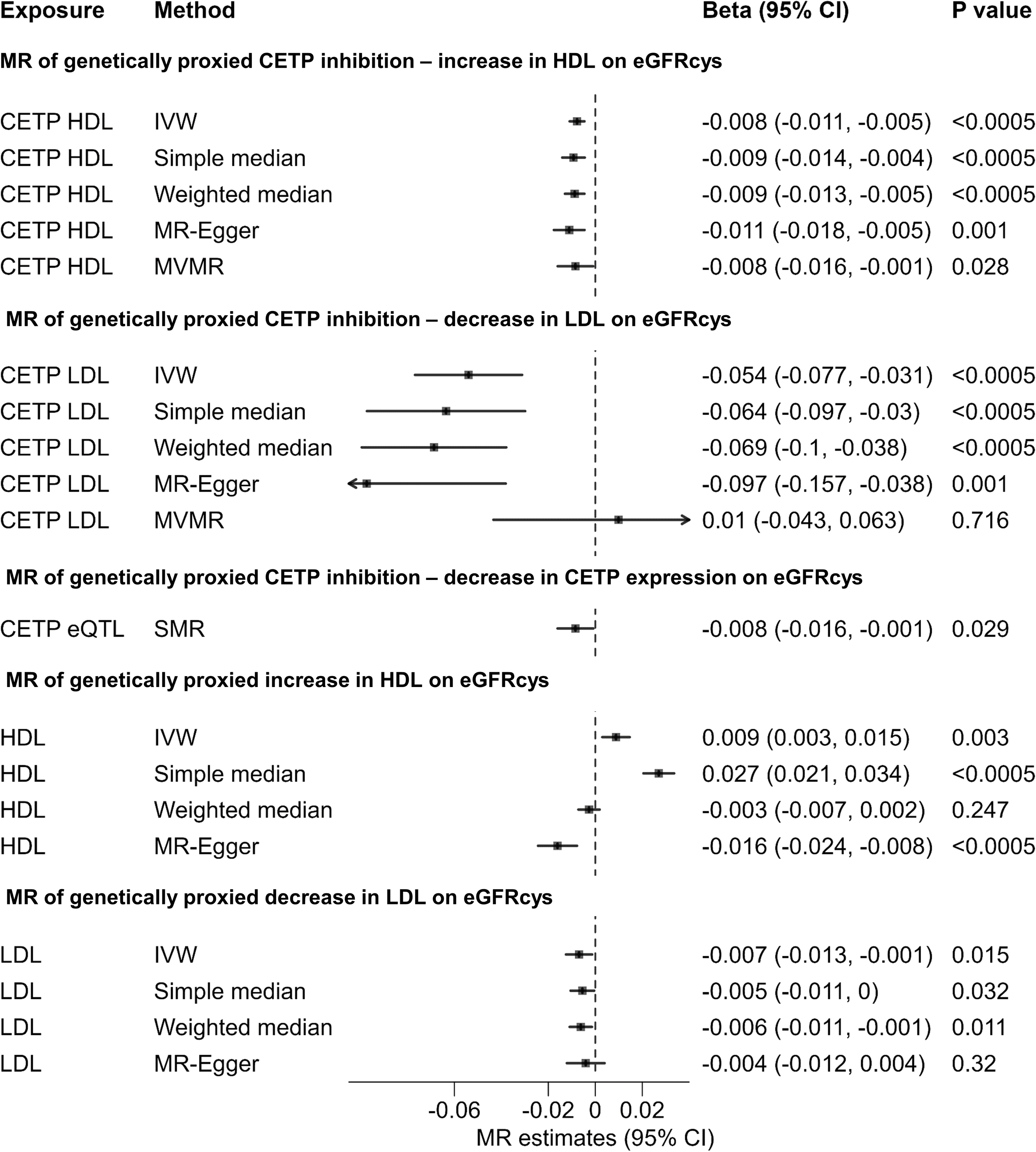
Mendelian randomization association of genetically predicted cholesteryl ester transfer protein inhibition, high-density lipoprotein, and low-density lipoprotein levels with serum cystatin C-estimated glomerular filtration rate. CETP indicates cholesteryl ester transfer protein; LDL, low-density lipoprotein; HDL, high-density lipoprotein; eGFRcys, serum cystatin C-estimated glomerular filtration rate; MR, Mendelian randomization; SMR, summary-based Mendelian randomization; MVMR, multivariable Mendelian randomization; IVW, inverse-variant weighted ; 95% CI, 95% confidence interval.

**Figure 3.**
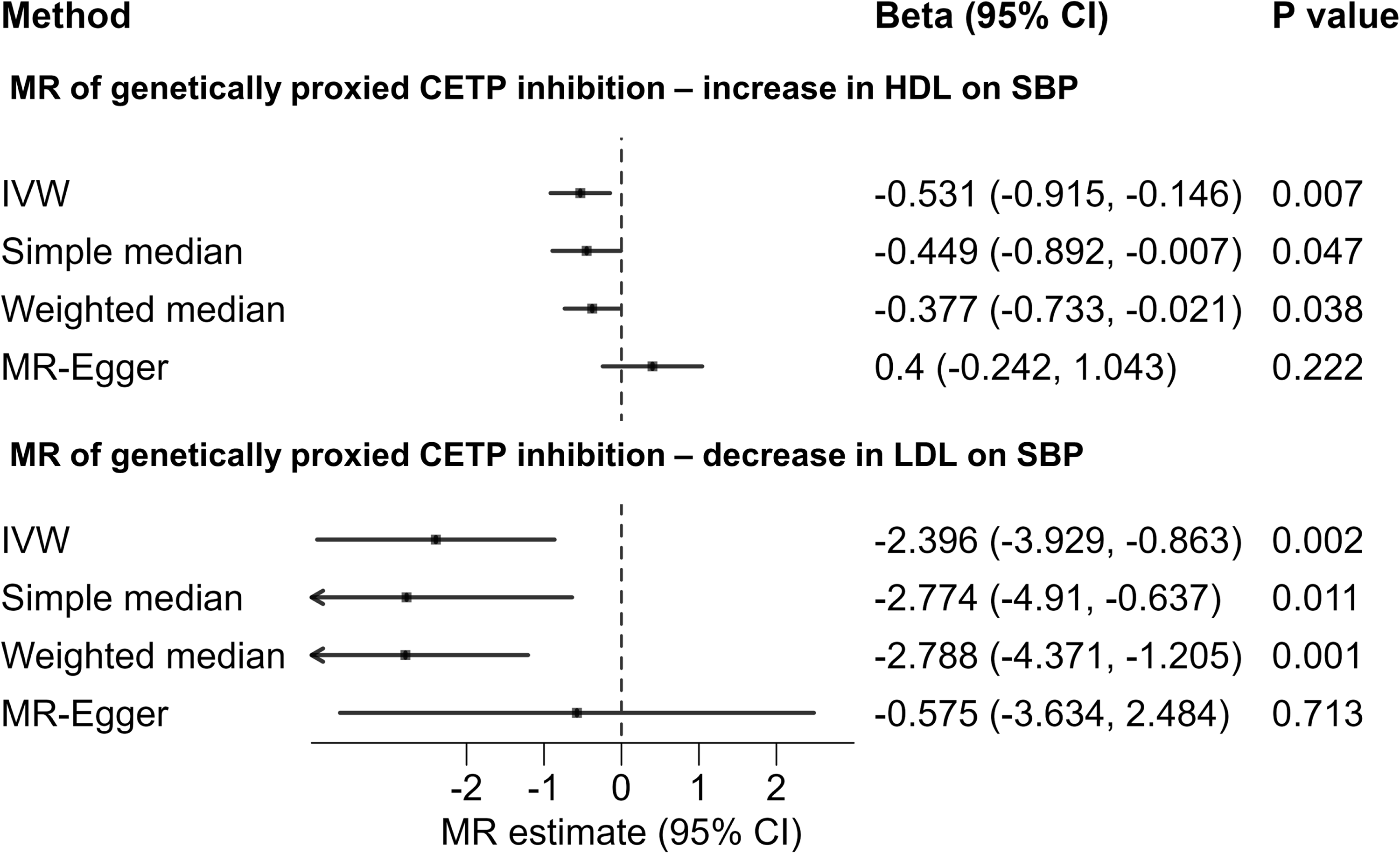
Mendelian randomization association of genetically predicted cholesteryl ester transfer protein inhibition with systolic blood pressure. CETP indicates cholesteryl ester transfer protein; LDL, low-density lipoprotein; HDL, high-density lipoprotein; SBP, systolic blood pressure; MR, Mendelian randomization; IVW, inverse-variant weighted ; 95% CI, 95% confidence interval.

When considering the genetic association with LDL as the exposure, genetically proxied CETP inhibition was associated with lower eGFRcys (effect size per 1 SD decrease in LDL level, −0.054, 95% CI −0.077, −0.031; p = 3.52 × 10^−06^) (**Figure 2, Figure S1, Table S3**). The estimates from the simple median, the weighted median and MR-Egger were consistent with the results from the main analysis; no evidence of directional pleiotropy was detected in the MR-Egger intercept (intercept p-value 0.12) (**Table S3)**. Genetically proxied CETP inhibition (decrease in LDL level as exposure) was not significantly associated with eGFRcrea, BUN, uACR, and MA risk, but associated with a reduced risk of CKD (OR per 1-SD decrease in LDL level, 0.449, 95% CI 0.278, 0.724; p = 0.001) (**Table S3, Figure S2-6)** and with SBP decrease (effect size per 1 SD decrease in LDL level, −2.34, 95% CI −3.93, −0.86, p = 0.002) (**Figure 3, Table S3)**.

Results from MVMR analysis showed that after genetically adjusting for LDL and SBP genetically proxied CETP inhibition (increased HDL level as exposure) was still associated with lower eGFRcys (−0.008, 95% CI −0.016, −0.001, p = 0.03) (**Table S4, Figure 2).** In contrast, when conducting MVMR using CKD as the outcome, only SBP exhibited a significant association with the risk of CKD after adjustments were made for genetically proxied HDL and LDL levels (**Table S5, Figure S6**).

#### Mendelian randomization of genetically proxied LDL and HDL

Across the genome, 498 uncorrelated SNPs (r^2^ < 0.001) were identified as genetic instruments for HDL, while 378 uncorrelated SNPs were identified for LDL. All of the SNPs had F-statistics greater than 10. The total variance explained by the genetic instruments is 6.7% and 6.24% for HDL and LDL, respectively **(Table S6-7)**.

IVW MR analysis identified a significant association of genetically increased HDL level with higher eGFRcys (effect size per 1-SD increase in HDL, 0.009, 95% CI 0.003, 0.015; p = 0.003) (**Figure 2, Figure S1**). MR estimate from simple median was consistent with the IVW estimate. However, the weighted median and MR Egger indicated a causal association in the opposite direction (MR Egger, effect size per 1-SD increase in HDL, −0.016, 95% CI −0.024, −0.008; p = 0.0002) (**Figure 2, Table S8**). The MR-Egger intercept test found evidence of directional pleiotropy (intercept p-value 4.22 × 10^−15^) (**Table S8)**. We obtained similar MR estimates when excluding SNPs within 1MB of the CETP gene **(Figure S1, Table S9)**.

Genetically increased HDL level was nominally significantly associated with lower BUN (−0.006, 95% CI, −0.011, −0.002), lower uACR (−0.023, 95% CI −0.041, −0.006), decreased CKD risk (OR, 0.916, 95% CI, 0.862, 0.974), but not with eGFRcrea. Genetically increased HDL level was significantly associated with decreased MA risk (OR, 0.918, 95% CI, 0.876, 0.963) and decreased SBP (−0.863, 95% CI, −1.313, −0.413) (**Table S8, Figure S2-6)**.

Genetically proxied decrease in LDL level was nominally significantly associated with lower eGFRcys (effect size per 1-SD decrease in LDL, −0.007, 95% CI −0.013, - 0.001; p = 0.015) **(Figure 2, Figure S1)**. The MR estimates obtained from the sensitivity analysis were consistent with the IVW MR estimate. The MR-Egger intercept test found no evidence of directional pleiotropy (intercept p-value 0.33) (**Table S10**). Similar MR estimates were obtained when excluding SNPs within 1MB of the CETP gene **(Figure S1, Table S11)**. No association was evident for SBP or CKD. Genetically proxied decrease in LDL level was significantly associated with lower eGFRcrea (effect size, −0.008, 95% CI −0.011, −0.003), lower uACR (−0.04, 95% CI, −0.058, −0.022), and decreased MA risk (OR, 0.919, 95% CI, 0.878, 0.961) (Table S10, Figure S2-6).

### Summary-based Mendelian Randomization

An inverse relationship was observed between CETP gene expression in blood and levels of LDL and HDL, as expected. A 1-SD increase in CETP gene expression was associated with an increase in LDL level (SD change, 0.23, 95% CI 0.19, 0.26) and a decrease in HDL level (SD change, −1.48, 95% CI −1.66, −1.30)) (**Table S12)**. Decreased CETP expression in blood was associated with lower eGFRcys (effect size per 1-SD decrease in gene expression, −0.008, 95% CI −0.016, −0.001; p_SMR_ = 0.029, p_HEIDI_ = 0.03), a reduced risk of CKD (OR per 1-SD decrease in gene expression, 0.85, 95% CI, 0.74, 0.98, p_SMR_ = 0.03; p_HEIDI_ = 0.63), and lower SBP (effect size, −0.71, 95% CI −1.177, −0.244, p_SMR_ = 0.003, p_HEIDI_ = 0.113) (**Figure 2, Table S12**). No significant association was observed between CETP gene expression and other renal function traits (**Table S12**).

### Colocalisation analysis

Colocalisation analysis produced high posterior probability for distinct causal variants (model H_3_) for pairwise combination of CETP gene expression and HDL levels reflecting regions of the gene showing concordant and discordant relationship between gene expression and HDL levels (**Figure S7**). Colocalisation analysis produced low posterior probabilities for both shared causal variant (model H_4_) and distinct causal variants (model H_3_) for the remaining pairwise combinations: CETP HDL level and eGFRcys, CETP gene expression and eGFRcys, CETP HDL level and CKD risk, CETP expression and CKD risk. (**Table S13, Figure S8).**

### Genetically predicted HMGCR inhibition

Twelve uncorrelated SNPs (r^2^ < 0.1) were identified as genetic proxies for HMGCR inhibition (**Table S14**). Genetically proxied HMGCR inhibition (reflected as an increase in LDL as exposure) had no significant association with eGFRcys (**Table S15, Figure S9)**, but associated with eGFRcrea (effect size per 1 SD decrease in LDL level, −0.01, 95% CI −0.016, −0.004; p = 0.002) (**Table S15**). 1-SD decrease in HMGCR gene expression in blood was associated with a decrease in LDL level, and a decrease in eGFRcys (effect size, −0.013, 95% CI −0.02, −0.005, p_SMR_ = 0.002, p_HEIDI_ = 0.402), and a decrease in eGFRcrea (−0.007, 95% CI −0.012, −0.002, p_SMR_ = 0.005, p_HEIDI_ = 0.435) (**Table S16, Figure S9**).

## 4. Discussion

Our results elucidate a complex relationship between HDL level, SBP and renal function. Through univariable and multivariable Mendelian randomization analyses, we demonstrate that increased HDL is specifically associated with reduced renal function defined by eGFRcys and this is independent of any SBP lowering effect. We do not observe an independent effect of LDL on eGFRcys. Counterintuitively, we find increased HDL is associated with a decreased risk of CKD in univariable MR analysis, which is consistent with previous MRs (17, 19, 46, 47). However, this association is not sustained in the MVMR analysis. Colocalisation analyses did not reveal either distinct or shared causal variants underpinning the genetically proxied CETP inhibition and decreased eGFRcys suggesting the relationship is HDL specific supporting the MR analysis using CETP variants that overcomes pleiotropic influences when using all HDL genetic variants from the whole genome. Furthermore, our results confirm the off-target effect of CETP inhibitors on blood pressure is related to the compound and not through its target effect on HDL.

Our finding that higher of HDL may cause kidney dysfunction is supported by several lines of biological and clinical evidence. Conventionally, the kidney’s role in HDL metabolism has been overlooked due to the assumption that HDL particles are too large to pass through the glomerular filtration barrier. However, emerging insights indicate that various components of HDL, such as ApoA-I and pre-β-HDL, do pass through this barrier due to their smaller molecular weight and shape characteristics (48). Furthermore, these components are not only filtered but also reabsorbed and metabolized by the kidney, suggesting a more complex interaction between HDL and renal function. Cholesterol loading and reduced ABCA1 expression were noted in podocytes when exposed to sera from diabetic patients with albuminuria as opposed to those without (49). A correlation was found between renal ABCA1 and ABCG1 expression and worsening of eGFR and diabetic nephropathy (50). APOL1, a component of specific HDL subfractions, has risk variants linked to nondiabetic CKD in African-Americans (51). A negative relationship was observed between total HDL cholesterol and eGFR, particularly among individuals with two APOL1 G1 risk variants (52). This suggests that APOL1 risk variants could adversely impact kidney function and modify HDL composition without changing total HDL levels (53).

Our results are in contrast with the majority of epidemiological studies shown that low levels of HDL cholesterol are a risk factor for renal dysfunction in both apparently healthy populations and those with existing chronic kidney disease (CKD). Meta-analyses have shown that lipid-lowering drugs like statins positively affect renal function, suggesting a potential cause-and-effect relationship between lipid levels and kidney health (54). However, the focus has primarily been on lowering overall cholesterol or LDL levels, not specifically on the effects of HDL. Our findings using genetic proxies for CETP HDL changes may reflect HDL specific effects. In epidemiological studies, reverse causality cannot be excluded as kidney dysfunction causes abnormalities in lipids and lipoproteins (55, 56).

Our study shows that genetically proxied HMGCR inhibition (with decreased LDL as exposure) was associated with reduced eGFRcrea, which is in concordance with previous MR (57). Previous studies on the renal implications of statins have yielded inconsistent findings. Some studies have indicated that statins could reduce proteinuria or decelerate the deterioration of kidney function (58–60). Whereas, other studies showed that high potency statins were associated with increased risk of acute kidney injury or interstitial nephritis (61, 62).

The strength of our study is the use of genetic proxies to investigate the effects of CETP inhibition on the outcomes. All of these variants had F statistics greater than 10, suggesting a low possibility for bias due to weak instruments. Simple median, weighted median and MR Egger methods were used in the sensitivity analysis to evaluate the robustness of the MR findings to potential pleiotropic effects of the instruments. One of the study limitations is that MR estimates reflect the lifelong effects of drug target perturbation and cannot be used to directly anticipate the effect magnitude from shorted pharmacological interventions. However, MR estimates remains a valuable indication of the existence and direction of causal effects (63). Moreover, the colocalization analysis gave low posterior probabilities for both shared causal variant scenario and distinct causal variants scenario, suggesting low statistical power to discriminate between these two scenario in the presence of MR evidence of a causal effect. A larger scale GWAS for eGFRcys may help to increase the statistical power. Another limitation is that the analyses of this study used genetic data derived from the individuals with European ethnicity, thereby limiting the generalizability of the results.

Our study has implications for therapeutic strategies targeting lipid modification for cardiovascular risk reduction. Careful consideration must be given to the potential renal effects of such interventions, especially in populations already at risk for kidney disease. Future studies should aim to clarify the mechanisms underlying the associations observed and should explore whether CETP inhibition has a direct impact on renal function beyond its effects on lipid levels.

## 5. Conclusion

In summary, our study provides valuable insights into the relationship between HDL and renal function. The results warrant validation of the nuanced roles of HDL and LDL on renal function and potential clinical implications of lipid management on renal outcomes.

## Data Availability

All data produced in the present study are available upon reasonable request to the authors

## 6. Acknowledgments

Conception and study design: SP, NNL. Data acquisition, analysis: NNL, TQBT. Drafting of manuscript: NNL. Critical revision of manuscript for important intellectual content: NNL, TQBT,DG, SP. Final approval of manuscript: All authors. Overall responsibility for the content as guarantors: SP, DG.

## 7. Sources of Funding

TQBT is supported by a British Heart Foundation MBPhD Studentship (FS/MBPhD/22/28005). SP is supported by the British Heart Foundation Centre of Excellence Award (RE/18/6/34217) and the United Kingdom Research and Innovation Strength in Places Fund (SIPF00007/1). DG is supported by the British Heart Foundation Centre of Research Excellence at Imperial College London. (RE/18/4/34215).

## 8. Disclosures

None.

## Notes

### Competing Interest Statement

The authors have declared no competing interest.

### Funding Statement

British Heart Foundation Centre of Excellence Award (RE/18/6/34217)
British Heart Foundation Centre of Research Excellence at Imperial College London. (RE/18/4/34215)

### Author Declarations

All data used in this study are publicly available. http://www.phenoscanner.medschl.cam.ac.uk/ https://www.mrbase.org/ https://gwas.mrcieu.ac.uk/ https://gtexportal.org/home/

